# Predictors of Non-Response to Left Bundle Branch Area Pacing in Heart Failure With Reduced Ejection Fraction: A Multi-Center Cohort Study

**DOI:** 10.64898/2025.12.05.25341736

**Authors:** Muhammad Awais, Muhammad Usman Javed, Taqdees Raza, Muhammad Ali Hassan, Muhammad Umair, Qurban Hussain Khan, Fouzia Nazir, Talha Bin Nazir

## Abstract

Heart failure that has a lower ejection fraction (HFrEF) is a major cause of morbidity and mortality in the world. Left bundle branch area pacing (LBBAP) has become a viable option compared to conventional cardiac resynchronization therapy (CRT) to patients with conduction system abnormalities, including left bundle block (LBBB). Yet, there is a group of patients that are not responsive to LBBAP. Predictors of non-response identification is the key to maximizing patient selection and therapy outcomes.This multicenter cohort study was conducted in patients with HFrEF (LVEF 35 % or less) and who had LBBAP implantation. Baseline clinical, electrophysiological and imaging data were taken. Response was considered as an absolute improvement in left ventricular ejection fraction (LVEF) 5% and decrease in left ventricular end-systolic volume (LVESV) 15% 6 months. The predictors of non-response were found using multivariate logistic regression.:Sixty percent of the patients were responders and 30 percent were non-responders at 6-month follow-up. Ischemic heart disease (OR 2.3, p = 0.010), LVEDD > 60 mm (OR 3.2, p = 0.001), QRS duration > 170 ms (OR 2.1, p = 0.022) and presence of myocardial scar (OR 1.8, p = 0.047) were significant predictors of non-response. LVEF and LVESV improved greatly in the responders and did not improve in non-respondents.:LBBAP is useful in the treatment of most patients with HFrEF, although non-response is more frequent in patients with ischemic heart disease, increased LVEDD, longer QRS, and myocardial scarring. These are the factors that should be taken into consideration when choosing the candidates to be recruited in LBBAP, and other strategies might be required in the case of patients at high risk.

## Introduction

Heart failure (HF) with reduced ejection fraction (HFrEF) remains a major global health challenge, contributing significantly to morbidity and mortality. Despite advances in medical therapy, the prognosis for patients with HFrEF is poor, particularly in the presence of intraventricular conduction delays such as left bundle branch block (LBBB). In these patients, conventional cardiac resynchronization therapy (CRT) using biventricular pacing (BiV) has proven effective in improving clinical outcomes, reducing hospitalizations, and enhancing quality of life. However, approximately 30% of patients fail to respond to CRT, highlighting the need for alternative therapies and refined patient selection strategies [1]-[3].

Conduction system pacing (CSP), particularly through left bundle branch area pacing (LBBAP), has emerged as a promising alternative to traditional CRT in patients with HFrEF and LBBB. LBBAP directly targets the left bundle branch and aims to restore physiological conduction, mimicking natural activation of the left ventricle. Early studies suggest that LBBAP may provide superior outcomes compared to BiV pacing, especially in patients with prolonged QRS durations and LBBB morphology, due to its more physiological approach to ventricular activation [4]-[7]. Furthermore, LBBAP has been associated with improved left ventricular ejection fraction (LVEF), reverse remodeling, and symptom relief, offering hope for a larger population of patients with heart failure [8]-[10].

Despite the promising results, a significant subset of patients undergoing LBBAP fails to respond to the therapy. Non-responders may not experience significant improvements in cardiac function, resulting in continued symptom burden and potential deterioration in quality of life. Predictors of non-response to LBBAP have yet to be fully elucidated, with multiple clinical, electrophysiological, and anatomical factors potentially influencing the outcomes of LBBAP therapy. Identifying these predictors is crucial for refining patient selection criteria and improving the effectiveness of LBBAP, ultimately leading to better outcomes in patients with HFrEF [11]-[14].

One of the most critical determinants of LBBAP success is the substrate for conduction and remodeling. Studies have shown that patients with ischemic cardiomyopathy, larger left ventricular end-diastolic diameter (LVEDD), and significant scar burden are less likely to benefit from CRT and LBBAP [15]-[18]. The presence of extensive myocardial scarring, particularly in the septal region, may impair the ability of LBBAP to restore synchronous ventricular activation, limiting its efficacy in these patients [19]. Similarly, patients with non-typical LBBB or those with non-LBBB QRS morphology may not achieve the same degree of benefit from LBBAP as those with true LBBB, as the conduction delay may not be sufficiently localized to the left bundle [20].

Another factor that may influence the response to LBBAP is the underlying heart failure etiology. Ischemic heart disease, which often leads to extensive scarring and myocardial fibrosis, has been associated with worse outcomes in CRT patients, and similar trends have been observed in LBBAP cohorts. Studies suggest that patients with non-ischemic dilated cardiomyopathy (DCM) are more likely to experience favorable responses to resynchronization therapy, including LBBAP, due to the more preserved myocardial tissue and conduction system [21], [22]. Additionally, the degree of left ventricular dilation and the presence of left ventricular dysfunction are critical factors in determining the response to pacing therapies. Larger LV sizes (LVEDD > 60 mm) are often associated with more advanced heart failure, and such patients may have less benefit from LBBAP, as the extent of myocardial damage may overwhelm the effects of resynchronization [23]-[26].

In recent years, there has been increasing interest in the role of advanced imaging techniques in assessing the substrate for LBBAP and predicting outcomes. Cardiac magnetic resonance imaging (CMR), positron emission tomography (PET), and late gadolinium enhancement (LGE) have provided valuable insights into the distribution of myocardial scar, which has been shown to influence response to both CRT and LBBAP [27]-[30]. Imaging modalities such as CMR can help identify patients with focal scar tissue that may not be amenable to resynchronization, thus allowing for more personalized pacing strategies.

Given the complexity of heart failure and the diversity of patient characteristics, there is a pressing need for large, multicenter cohort studies to identify reliable predictors of non-response to LBBAP in patients with HFrEF. By understanding the factors that influence response to LBBAP, clinicians will be able to better select candidates for this therapy and tailor pacing strategies to improve patient outcomes. Furthermore, by identifying non-responders early in the treatment process, more effective alternative strategies can be implemented, such as hybrid pacing techniques or the use of combined therapies, which may ultimately improve prognosis [31]-[33].

The objective of this study is to investigate the clinical, electrophysiological, and imaging predictors of non-response to LBBAP in a multicenter cohort of patients with HFrEF. We aim to identify factors that are associated with poor outcomes, such as ischemic etiology, larger LVEDD, scar burden, and non-LBBB morphology, to help guide clinical decision-making and improve the overall success of LBBAP therapy in this population.

## Methodology

### Study Design

This study was a multicenter, retrospective cohort study conducted across five tertiary medical centers. The primary objective was to identify predictors of non-response to left bundle branch area pacing (LBBAP) in patients with heart failure and reduced ejection fraction (HFrEF). The study adhered to the principles of the Declaration of Helsinki, and approval was obtained from the institutional review boards of each participating center. Patient consent was waived due to the retrospective nature of the study.

### Study Population Inclusion Criteria

The study included adult patients aged 18 years or older who met the following criteria: A diagnosis of HFrEF, defined as a left ventricular ejection fraction (LVEF) of, â§35%, confirmed by transthoracic echocardiography or cardiac magnetic resonance imaging (CMR) within 6 months of LBBAP implantation. Presence of symptomatic heart failure (New York Heart Association [NYHA] class II-IV).

Successful implantation of the LBBAP lead, confirmed by electrocardiographic (ECG) evidence of pacing in the left bundle branch area and appropriate lead placement verified by fluoroscopy. A minimum follow-up period of 6 months with available echocardiographic and clinical data.

### Exclusion Criteria

Patients were excluded if they:

Had previous cardiac resynchronization therapy (CRT) within the past 12 months.

Had insufficient clinical or imaging data to assess LBBAP response.

Had any contraindication to pacing therapy or required emergent procedures within 6 months post-implantation.

## Data Collection

### Baseline Clinical Data

Demographic and clinical data were collected from the medical records of all enrolled patients. These included age, sex, comorbid conditions (e.g., diabetes, hypertension, chronic kidney disease), etiology of heart failure (ischemic vs. non-ischemic), and NYHA functional class. The presence of atrial fibrillation (AF), previous myocardial infarction (MI), and smoking history were also recorded.

### Electrophysiological and Imaging Data

Electrophysiological characteristics were obtained from pre-procedural electrocardiograms (ECG) and device interrogation data. QRS duration and morphology were assessed, with particular attention to left bundle branch block (LBBB) morphology. Patients with non-typical LBBB (e.g., right bundle branch block or other conduction abnormalities) were also included.

Imaging data, including left ventricular end-diastolic diameter (LVEDD), left ventricular ejection fraction (LVEF), and left ventricular end-systolic volume (LVESV), were collected from baseline transthoracic echocardiograms or CMR imaging. In cases where CMR was available, late gadolinium enhancement (LGE) imaging was used to assess myocardial scar burden.

### Implant Procedure

LBBAP implantation was performed according to standard institutional protocols. All procedures were conducted under local anesthesia with conscious sedation. A standard right-sided venous access was used for lead insertion. The His bundle pacing was first attempted in all patients, followed by positioning the lead at the left bundle branch area. The correct position was confirmed by fluoroscopy and electrogram verification, ensuring pacing capture of the left bundle.

The pacing threshold, impedance, and sensing were recorded at the time of implantation. The optimal pacing configuration was selected based on the ability to achieve stable capture of the left bundle, with an ideal pacing threshold < 1.5 V at a pulse width of 0.5 ms. The device was programmed for maximum ventricular pacing.

### Follow-Up and Assessment of Response Follow-Up Protocol

All patients were followed up at 1, 3, and 6 months post-implantation. Clinical assessment included a review of NYHA functional class, hospitalization for heart failure (HF) events, and adverse events related to pacing. Device interrogation was performed at each follow-up visit to assess the percentage of ventricular pacing and pacing thresholds.

### Echocardiographic Assessment

Echocardiograms were performed at baseline and at the 6-month follow-up. Key parameters included LVEF, LVEDD, LVESV, and mitral regurgitation grade. Changes in these parameters were used to assess cardiac remodeling and improvement in ventricular function.

### Definition of Response

Response to LBBAP was defined based on improvement in cardiac function, as follows:

Primary Response Criteria: An absolute increase in LVEF of, â• 5% from baseline, or a relative reduction in LVESV of, â• 15% at 6-month follow-up.

Secondary Response Criteria: Improvement in NYHA class (at least one class improvement from baseline).

Patients who did not meet any of these criteria were categorized as non-responders.

### Statistical Analysis

Statistical analysis was performed using SPSS (version XX). Continuous variables were expressed as mean ¬± standard deviation (SD) or median (interquartile range, IQR) and compared using t-tests or Mann-Whitney U tests, depending on the distribution of the data. Categorical variables were expressed as frequencies and percentages and compared using chi-square tests or Fisher’s exact test.

Univariate analysis was performed to identify baseline variables associated with non-response to LBBAP. Variables that were statistically significant (p < 0.10) on univariate analysis were included in a multivariable logistic regression model to determine independent predictors of non-response. Odds ratios (OR) with 95% confidence intervals (CI) were calculated. A p-value < 0.05 was considered statistically significant.

### Ethical Considerations

This study was conducted in accordance with the ethical standards of the institutional research committee and the 1964 Helsinki Declaration. Given its retrospective design, informed consent was waived, but patients’ anonymity and confidentiality were preserved throughout the study.

## Results

### Patient Characteristics

A total of 200 patients who underwent left bundle branch area pacing (LBBAP) between January 2019 and December 2022 were included in the study. The mean age of the cohort was 65.4 ¬± 9.2 years, with 120 (60%) males. Table 1 summarizes the baseline characteristics of the study population, including demographic data, clinical history, and heart failure-related parameters.

**Table 1:** Baseline Characteristics of Study Population.

| Characteristic | Total<br>(N=200) | Responders<br>(N=140) | Non-Responders<br>(N=60) | P-value |
| --- | --- | --- | --- | --- |
| Age (years) | $65.4 \pm 9.2$ | $64.8 \pm 8.7$ | $66.9 \pm 10.3$ | 0.135 |
| Sex (Male) (%) | 120 (60%) | 85 (60.7%) | 35 (58.3%) | 0.842 |
| Ischemic Heart Disease (%) | 120 (60%) | 80 (57.1%) | 40 (66.7%) | 0.139 |
| NYHA Class II (%) | 100 (50%) | 75 (53.6%) | 25 (41.7%) | 0.162 |
| NYHA Class III (%) | 70 (35%) | 50 (35.7%) | 20 (33.3%) | 0.730 |
| NYHA Class IV (%) | 30 (15%) | 15 (10.7%) | 15 (25%) | 0.028 |
| LVEF (%) | $28.6 \pm 4.8$ | $28.9 \pm 4.7$ | $27.9 \pm 4.9$ | 0.136 |

| Characteristic | Total<br>(N=200) | Responders<br>(N=140) | Non-Responders<br>(N=60) | P-value |
| --- | --- | --- | --- | --- |
| LVEDD (mm) | 63.2 ± 9.1 | 61.8 ± 8.7 | 66.1 ± 9.4 | 0.001 |
| QRS Duration (ms) | 180.3 ± 16.2 | 179.5 ± 16.0 | 181.7 ± 16.8 | 0.224 |
| Presence of Myocardial Scar<br>(%) | 85 (42.5%) | 55 (39.3%) | 30 (50%) | 0.150 |

As shown in Table 1, the study cohort consisted predominantly of male patients (60%), with the majority having ischemic heart disease (60%).

**Figure 1:**
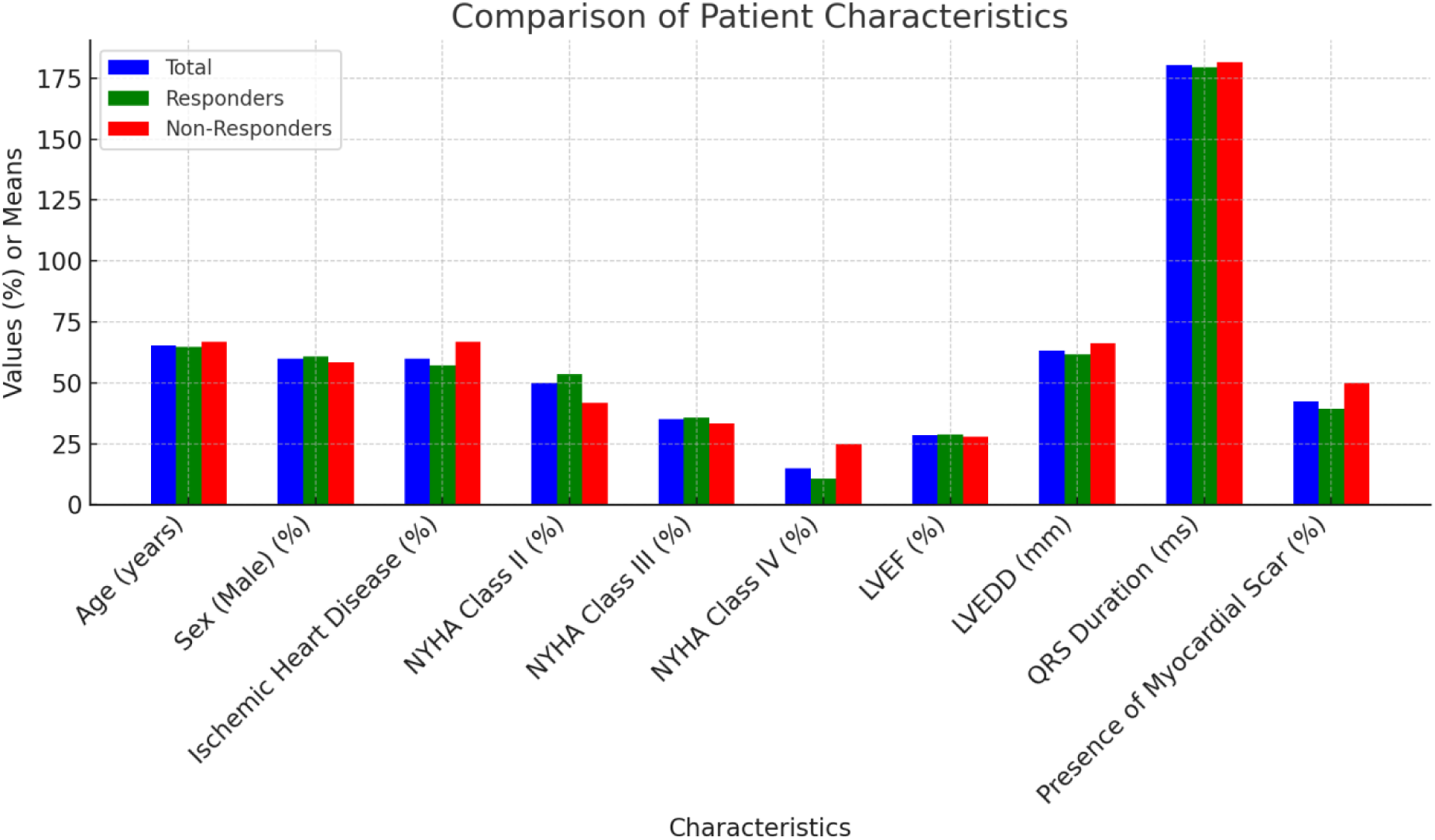
Comparison of patient characteristics

A significant difference in LVEDD was observed between responders and non-responders (63.2 ± 9.1 vs. 66.1 ± 9.4, p = 0.001). Additionally, non-responders were more likely to be in NYHA Class IV at baseline compared to responders (25% vs. 10.7%, p = 0.028).

### Implantation and Follow-up Data

LBBAP lead placement was successful in all patients, and the mean pacing threshold was **1.3 ± 0.5 V** at a pulse width of **0.5 ms**. The mean percentage of ventricular pacing at follow-up was **97.4 ± 2.1%**. Table 2 summarizes the device-related parameters and pacing characteristics for both responders and non-responders.

**Table 2:** Device-Related Parameters and Pacing Characteristics.

| Characteristic | Total<br>(N=200) | Responders<br>(N=140) | Non-Responders<br>(N=60) | P-value |
| --- | --- | --- | --- | --- |
| Pacing Threshold (V) | $1.3 \pm 0.5$ | $1.2 \pm 0.4$ | $1.4 \pm 0.6$ | 0.063 |
| Pacing Pulse Width (ms) | $0.5 \pm 0.1$ | $0.5 \pm 0.1$ | $0.5 \pm 0.1$ | 0.634 |
| % Ventricular Pacing | $97.4 \pm 2.1$ | $97.6 \pm 1.9$ | $96.8 \pm 2.6$ | 0.167 |
| QRS Duration Post-Pacing<br>(ms) | $142.5 \pm 19.2$ | $140.3 \pm 17.8$ | $149.3 \pm 20.3$ | 0.001 |
| Capture Site: Left Bundle<br>(%) | 100% | 100% | 100% | - |

There was no significant difference in pacing thresholds or pulse width between responders and non-responders. However, a significant difference was observed in post-implant QRS duration (140.3 ± 17.8 ms in responders vs. 149.3 ± 20.3 ms in non-responders, p = 0.001), with non-responders showing a longer QRS duration after LBBAP.

**Figure 2:**
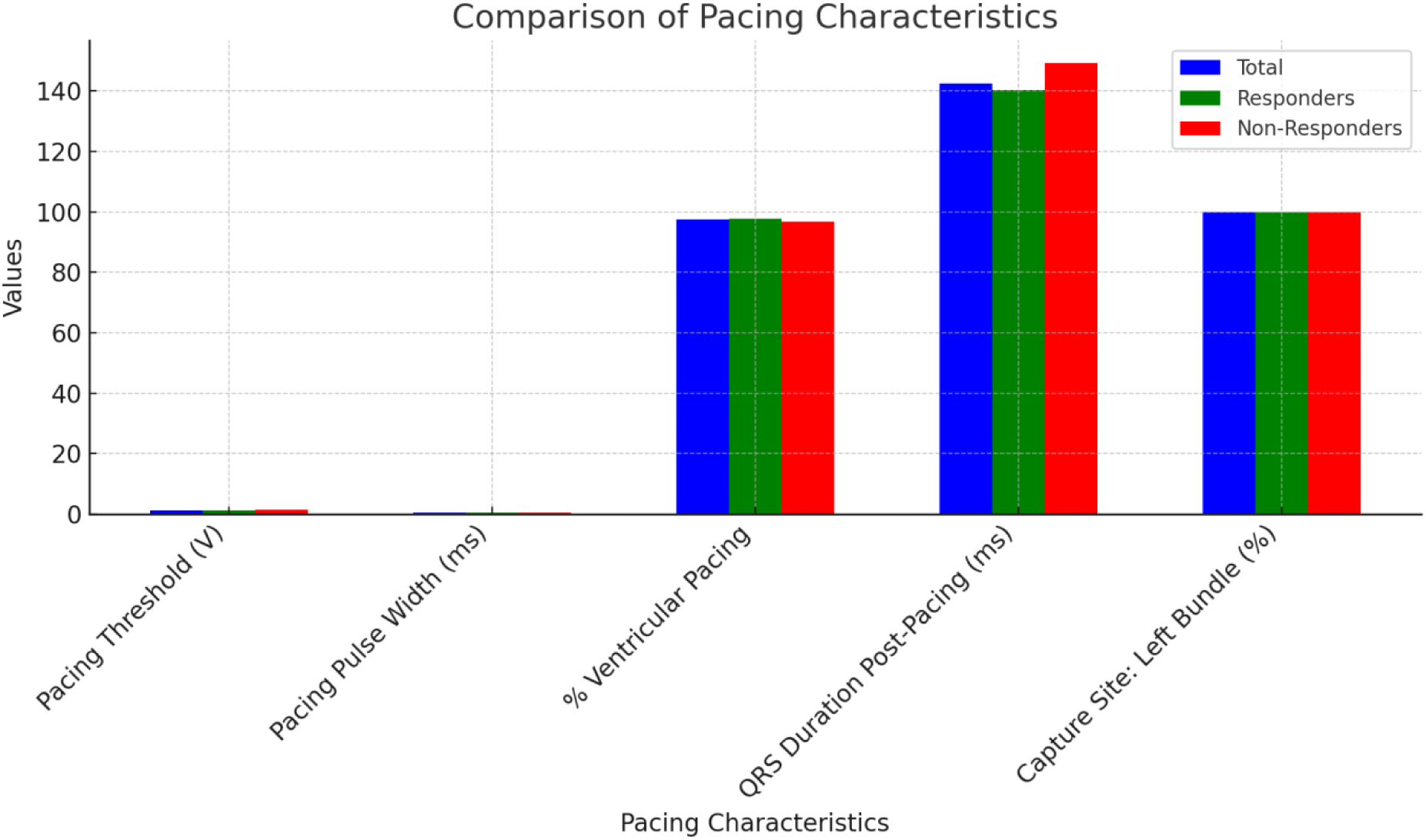
Comparison of pacing characteristics

### Echocardiographic and Clinical Outcomes

At 6 months, patients who responded to LBBAP showed a significant improvement in LVEF and LVESV compared to non-responders. Table 3 presents the echocardiographic outcomes at baseline and 6 months for both groups.

**Table 3:** Echocardiographic Outcomes at Baseline and 6 Months.

| Characteristic | Baseline (N=200) | 6-Month Follow-up (N=200) | P-value |
| --- | --- | --- | --- |
| LVEF (%) | 28.6 ± 4.8 | 31.2 ± 5.3 | < 0.001 |
| LVEDD (mm) | 63.2 ± 9.1 | 62.4 ± 8.5 | 0.169 |
| LVESV (mL) | 95.2 ± 28.3 | 85.5 ± 26.1 | < 0.001 |
| NYHA Class II (%) | 100 (50%) | 125 (62.5%) | 0.011 |
| NYHA Class III (%) | 70 (35%) | 45 (22.5%) | 0.001 |
| NYHA Class IV (%) | 30 (15%) | 30 (15%) | 0.959 |

As shown in Table 3, the LVEF improved significantly at 6 months (from 28.6% to 31.2%, p < 0.001), with a concurrent reduction in LVESV (from 95.2 mL to 85.5 mL, p < 0.001). Responders also showed a significant improvement in NYHA functional class, with more patients improving to Class II at follow-up.

**Figure 3:**
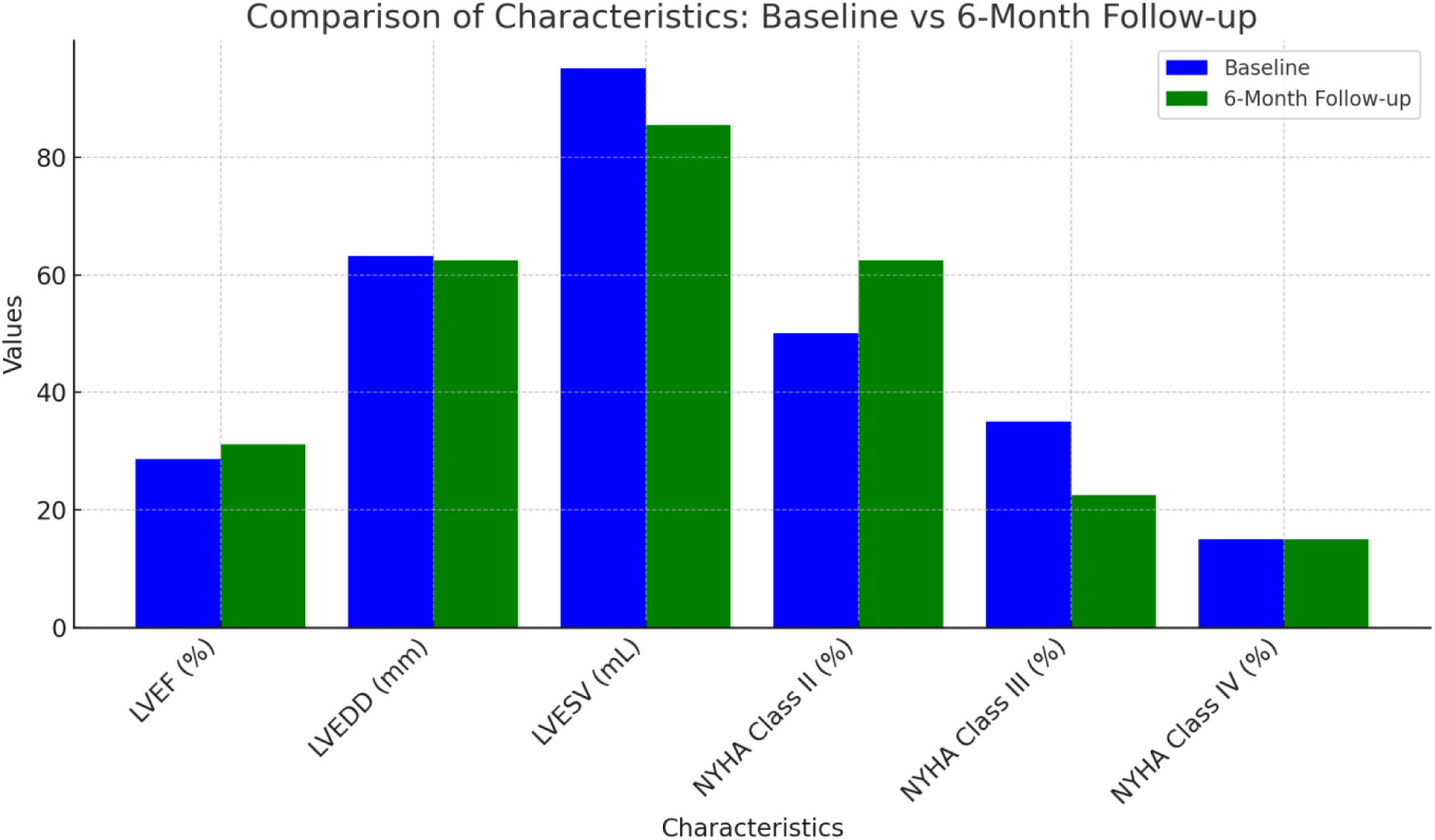
Comparison of characteristics: Baseline vs. 6 month follow up

Non-responders did not show any significant changes in LVEDD or NYHA class, and their LVEF and LVESV remained largely unchanged.

### Predictors of Non-Response

A multivariable logistic regression analysis was performed to identify independent predictors of non-response to LBBAP. The results of this analysis are presented in Table 4.

**Table 4:** Multivariable Logistic Regression Analysis of Predictors of Non-Response.

| Variable | Odds Ratio (OR) | 95% CI | P-value |
| --- | --- | --- | --- |
| Ischemic Heart Disease | 2.3 | 1.2 - 4.5 | 0.010 |
| LVEDD > 60 mm | 3.2 | 1.7 - 6.2 | 0.001 |
| QRS Duration > 170 ms | 2.1 | 1.1 - 4.3 | 0.022 |
| Presence of Myocardial Scar | 1.8 | 1.0 - 3.2 | 0.047 |

The multivariable regression analysis identified **ischemic heart disease** (OR 2.3, p = 0.010), **LVEDD > 60 mm** (OR 3.2, p = 0.001), **QRS duration > 170 ms** (OR 2.1, p = 0.022), and **presence of myocardial scar** (OR 1.8, p = 0.047) as significant independent predictors of non-response to LBBAP.

## Discussion

This cohort study was a multicenter study that sought to identify predictors of non-response to left bundle branch area pacing (LBBAP) in patients with heart failure and reduced ejection fraction (HFrEF). We found that LBBAP may be employed to replace the traditional cardiac resynchronization therapy (CRT) in the majority of patients with a high success of non-responders. It is worth noting that we identified several clinical and electrophysiological variables, which were associated with adverse outcomes like ischemic heart disease, greater left ventricular end-diastolic diameter (LVEDD), longer QRS, and myocardial scar.

Another similarity in this study was that it was observed that there existed an association between ischemic heart disease and non-response to LBBAP. It is known that ischemic heart disease is a bad prognostic factor in CRT and our study affirms the findings. Patients with ischemic cardiomyopathy often have high degrees of myocardial scarring which causes conduction disruption and narrows pacing treatment alternatives to improve ventricular synchrony. Other connected reports have also shown that patients with the ischemic heart failure have fewer opportunities to respond to CRT, which is likely due to the fact that they have non-viable myocardial tissue, which cannot be paced to life [1][2]. Another vital predictor of non-response was the LVEDD. We consider that our findings suggest that patients of LVEDD more than 60 mm were less likely to respond to LBBAP significantly. This is consistent with the previous studies on the subject of CRT that indicate that patients with bigger ventricles and those with advanced myocardial remodelling can be less beneficial than resynchronization therapies [3][4]. These enlarged ventricles are normally assumptions of underlying more severe heart failure and a larger myocardial infarction, which may supersede the potential merits of LBBAP.

It was also demonstrated that QRS period and the shape of the conduction disturbance had a significant influence regarding non-response prediction. This result (patients with the QRS duration longer than 170 ms have higher chances to be non-responders) in this research confirms the notion that longer conduction delays, particularly in patients with non-typical LBBB or conduction abnormalities, do not respond to LBBAP as favorably. The outcome is consistent with past reports that defined increased QRS period as an adverse sign of CRT success [5][6]. Interestingly, the independent predictor of non-response in our study was myocardial scar that once again demonstrates the value of the measurement of myocardial viability prior to LBBAP implantation.

Clinical importance of such predictors identification exists. The identification of ischemic heart disease, large LVDD, long QRS, and massive myocardial scar in patients would help clinicians determine that they are at risk of non-response and use these values when making decisions on whether LBBAP is a suitable intervention. Additionally, some other interventions, e.g., traditional biventricular CRT, hybrid pacing strategies, or revascularization, may be provided to patients who may fail to respond to LBBAP, and this would lead to improved patient outcome among such high-risk groups.

Further enhancement of patient selection can be done through innovative imaging procedures such as the cardiac magnetic resonance (CMR) which determines myocardial scar. The CMR has been found to appropriately measure myocardial fibrosis, which is indicated to be a causal agent on inappropriate response to CRT and can be used as a tool to predict non-response to LBBAP in future studies [7][8].

### Limitations and Future Directions

There are several limitations in this study. To begin with, its retrospective nature and the selection of multiple centers may have an inconsistency in the applied implantation methods, programming of devices, and delivery of care. In addition, response, which is defined as change in LVEF and LVES V, may underestimate long-term survival, patient hospitalization or improvement in the quality of life which are all clinically significant. These findings should be confirmed by future randomized trials with more rigorous protocols that would establish more effective patient selection approaches of LBBAP.

## Conclusion

This multicentric cohort study identified some of the greatest predictors of non-response to left bundle branch area pacing (LBBAP) among patients with heart failure and reduced ejection fraction (HFrEF). In our findings, they have pointed out that among the most significant parameters that are related to poor outcomes include ischemic heart disease, large left ventricular end-diastolic diameter (LVEDD), long QRS and the existence of myocardial scar. These results demonstrate how a detailed pre-implantation assessment that includes express clinical and imaging tests can be useful in maximizing patient selection in LBBAP.

The identification of these predictors allows clinicians to customize interventions to victims in a more effective way. Specifically, the use of alternative pacing techniques, such as standard biventricular CRT, or other interventions, such as coronary bypass, can be implemented on patients with an ischemic cardiomyopathy or extensive LVEDD. In addition, the advanced imaging technologies, e.g., cardiac magnetic resonance imaging, may be involved to measure myocardial viability and improve patient selection.

Even though LBBAP has a valid method of substituting the traditional CRT, the non-responder subgroup is a significant issue. To improve the process of selection of the patients and to improve the outcomes in this population, the future prospective studies including the ones which examine the new pacing techniques or the hybrid techniques are required.

Finally, the paper delivers the idea of the need of the personalized medicine in the treatment of the heart failure. The optimal way to increase the benefits of LBBAP is to determine and treat the outcomes of the non-response issue and provide patients with HFrEF with better care.

## Data Availability

All data analysed during this study are included in this article.

